# Humoral Response Dynamics Following Infection with SARS-CoV-2

**DOI:** 10.1101/2020.07.16.20155663

**Authors:** Louis Grandjean, Anja Saso, Arturo Torres, Tanya Lam, James Hatcher, Rosie Thistlethwayte, Mark Harris, Timothy Best, Marina Johnson, Helen Wagstaffe, Elizabeth Ralph, Annabelle Mai, Caroline Colijn, Judith Breuer, Matthew Buckland, Kimberly Gilmour, David Goldblatt, the Co-Stars Study Team

**Author notes:** **Corresponding author contact details** Dr Louis Grandjean, Department of Infection, Inflammation and Immunity, Institute of Child Health, 30 Guilford Street, London, WC1N. Contributed equally (LG is also corresponding author). Contributed equally (Arturo Torres, Tanya Lam). The Co-Stars Study Team: Dorcas Mirambe-Korsah, Fernanda Torrente, Jakub Wyszynski Victoria Gander, Amy Leonard, Louise Myers, Aimee Vallot, Camille Paillas, Rose Fitzgerald, Adam Twigg, Rabia Manaf, Lois Gibbons, Hollie Powell, Richard Nar-Dorh, Ally Gray, Elias Fernandez, Aline Minja, Emily Beech, Waffa Girshab, Pei Shi Chia, Kate Webb, Malti Nakrani, Kim Gardiner, Valerija Karaluka, Karen Ryan, Dorothy Lee, Katie Groves, Hamad Khan, Shamime Nsubuga, Olivia Rosie-Wilkinson, Julia Spires, Nuria Sanchez-Clemente, Sapriya Kaur, Natasha Carroll□, Jemma Efford, Gabriel Bredin, Celma Marisa Dos Santos Domingues□, Sophie Foxall□, Helen Ashton, Abbey Afzal, Sally Mainland, Kate Crumpler, Lucinda Dawson, Claire Smith, Maria Tabbu, Laura Chiverton, Jade Sugars, Jordan Mooney, Dorothy Chikusu, Fariba Tahami, Baratth Samy, Shomona Begum, Dhimple Patel, Philippa Wiltshire, Annie Susay, Anna Ryan, Luke Lancaster, Kavita Thind, Kate Speller, Rachel Sterling, Connor Tugulu, Sandhya Ghurburrun, Steffi Gray, Joy Mugas, Moe Kishma, Kathleen Akpokomua, Sophie White, Eleana Pieri, Sabina Shamsad, Demi Alexandrou, Odera Aguele, Katherine Miles, Anamika Jain, Subishma Gautam, Oliver Simms. **Author Contributions** The first draft of the manuscript was prepared by: LG, AS, AT, TL, JH, RT, MH, TB, MJ, HW, ER, AM, CC, JB, KG, DG. The study was designed and implemented by: LG, AS, AT, TL, JH, RT, MH, TB, MJ, HW, ER, AM, CC, JB, KG, DG & the Co-Stars Team. Testing for SARS-CoV-2 RNA and antibody was conducted by: LG, AS, AT, TL, JH, RT, MH, TB, MJ, HW, ER, AM, CC, JB, KG, DG.

## Abstract

**Introduction:** Severe Acute Respiratory Syndrome Coronavirus-2 (SARS-CoV-2) specific antibodies have been shown to neutralize the virus in-vitro. Understanding antibody dynamics following SARS-CoV-2 infection is therefore crucial. Sensitive measurement of SARS-CoV-2 antibodies is also vital for large seroprevalence surveys which inform government policies and public health interventions. However, rapidly waning antibodies following SARS-CoV-2 infection could jeopardize the sensitivity of serological testing on which these surveys depend.

**Methods:** This prospective cohort study of SARS-CoV-2 humoral dynamics in a central London hospital analyzed 137 serial samples collected from 67 participants seropositive to SARS-CoV-2 by the Meso-Scale Discovery assay. Antibody titers were quantified to the SARS-CoV-2 nucleoprotein (N), spike (S-)protein and the receptor-binding-domain (RBD) of the S-protein. Titers were log-transformed and a multivariate log-linear model with time-since-infection and clinical variables was fitted by Bayesian methods.

**Results:** The mean estimated half-life of the N-antibody was 52 days (95% CI 42-65). The S- and RBD-antibody had significantly longer mean half-lives of 81 days (95% CI 61-111) and 83 days (95% CI 55-137) respectively. An ACE-2-receptor competition assay demonstrated significant correlation between the S and RBD-antibody titers and ACE2-receptor blocking in-vitro. The time-to-a-negative N-antibody test for 50% of the seropositive population was predicted to be 195 days (95% CI 163-236).

**Discussion:** After SARS-CoV-2 infection, the predicted half-life of N-antibody was 52 days with 50% of seropositive participants becoming seronegative to this antibody at 195 days. Widely used serological tests that depend on the N-antibody will therefore significantly underestimate the prevalence of infection following the majority of infections.

**Significance statement:** We believe that our study has significant and urgent public health and translational impact. Firstly, our findings demonstrate that the half-life of the SARS-CoV-2 nucleoprotein antibody is only 52 days. This has immediate and important implications for large-scale seroprevalence surveys, government policy and mathematical modelling predictions which rely on serological tests that target this antibody. Secondly, the slower decay of the SARS-CoV-2 spike protein antibody identified in this study makes assays to the spike protein a more reliable target for serological assays in the longer term. We demonstrate a strong positive linear correlation between spike/RBD antibody and ACE-2 receptor binding in vitro. Our findings are therefore likely to reflect the time to loss of a functional antibody response in SARS-CoV-2.

**Funding:** GOSH charity, Wellcome Trust (201470/Z/16/Z and 220565/Z/20/Z). GOSH NIHR Funded Biomedical Research Centre.

**Trial registration number:** NCT04380896.

## Introduction

Since appearing as a cluster of pneumonia cases in December 2019 in Wuhan, China, Coronavirus disease (COVID-19) has rapidly spread worldwide.(1) As of July 7^th^, there have been 11,863,477 cases, resulting in 544,949 deaths and a global health crisis, with significant social, economic and public health implications.(2) COVID-19 is caused by severe acute respiratory syndrome coronavirus-2 (SARS-CoV-2), an enveloped RNA β-coronavirus.(3) Specific immunoglobulin (IgG) antibody responses to the SARS-CoV-2 trimeric spike (S) protein, nucleoprotein (N) protein and the receptor-binding domain (RBD) develop between 6-15 days following disease-onset.(4) The S-protein, which contains the RBD, binds to host cells via the angiotensin-converting-enzyme-2 (ACE-2) receptor, and membrane fusion occurs before viral entry.(5, 6) The N-protein plays an important role in transcription enhancement and viral assembly.(7)

The dynamics and duration of the antibody response to S-protein, N-protein and the RBD following infection or exposure with SARS-CoV-2 remain poorly understood.(8–10) Quantifying the humoral response shortly after infection limits the inferences that can be made about longer term dynamics. SARS-CoV-2-specific antibodies (particularly to the S-protein) detected in convalescent individuals have been shown to strongly correlate with virus-specific T-cell responses and viral neutralisation *in vitro*, as well as protection against disease in animal models, following passive transfer of selected monoclonal antibodies.(8, 11–15) It is unknown whether re-infection in humans occurs following primary infection with SARS-CoV-2. However, re-infection did not occur in rhesus macaques that were re-challenged in the presence of detectable endogenous antibodies.(16, 17) Taken together, these findings highlight the importance of characterising humoral dynamics following SARS-CoV-2 infection.

A large seroprevalence survey undertaken in Spain, recently reported a SARS-CoV-2 seroprevalence of 5.0%(18). This study used a chemiluminescent assay to detect nucleoprotein antibody together with a point of care lateral flow device. In contrast, Public Health England undertook a SARS-CoV-2 spike antibody seroprevalence survey of 1000 samples from blood donors in London estimating a seroprevalence of 17.5% in the first week of April(19). Whilst many socio-demographic factors could account for the different estimates of seroprevalence between these studies, one explanation may be the relative decay of the spike and the nucleoprotein antibodies.

Longitudinal serological data from Taiwan’s national SARS database estimated the half-life of SARS-CoV-1 neutralizing antibody to be 45 days,(20) whilst 74% of hospitalized patients in China had detectable SARS-CoV-1 IgG at 36 months post-infection.(21) After pauci-symptomatic MERS infection, antibodies rapidly decayed and were undetectable within one year.(22) However, following severe disease, antibodies have been detected up to 34 months post-infection.^23^

In order to evaluate SARS-CoV-2 antibody kinetics and longevity, we launched the Covid-19 Staff Testing of Antibody Responses Study (Co-STARS), a longitudinal prospective cohort study of healthcare workers designed to measure serial quantitative antibody levels over 1-year. In parallel with serological data, detailed demographic, clinical and socioeconomic data was also collected across different hospital departments, to provide a comprehensive insight into factors that may influence antibody dynamics.

## Results

### Socio-Demographic and Clinical Details

A total of 137 longitudinal samples that were in pairs or triplicate from 67 of the first participants in the Co-Stars study were available for analysis. Fifty-three participants (79%) of the cohort were women and 25% were from Black, Asian, Minority Ethnic (BAME) backgrounds (17/67). Importantly, none of the participants had been admitted to hospital and only 5% had underlying comorbidities. Age, clinical, and demographic variables for the cohort are reported in Supplementary Table 1.

### Model Fit

The combination of co-variates that provided the best model fit was determined by optimizing the Deviance Information Criterion (**Supplementary Table 2**). Log-linear models were fitted to the decay of each SARS-CoV-2 antibody with confidence limits on the slope included with and without individual random effects (**Figure 1**). Monte Carlo Markov Chain (MCMC) plots converged appropriately for the 3 antibodies and estimated parameters (**Supplementary Figure 3**).

**Figure 1.**
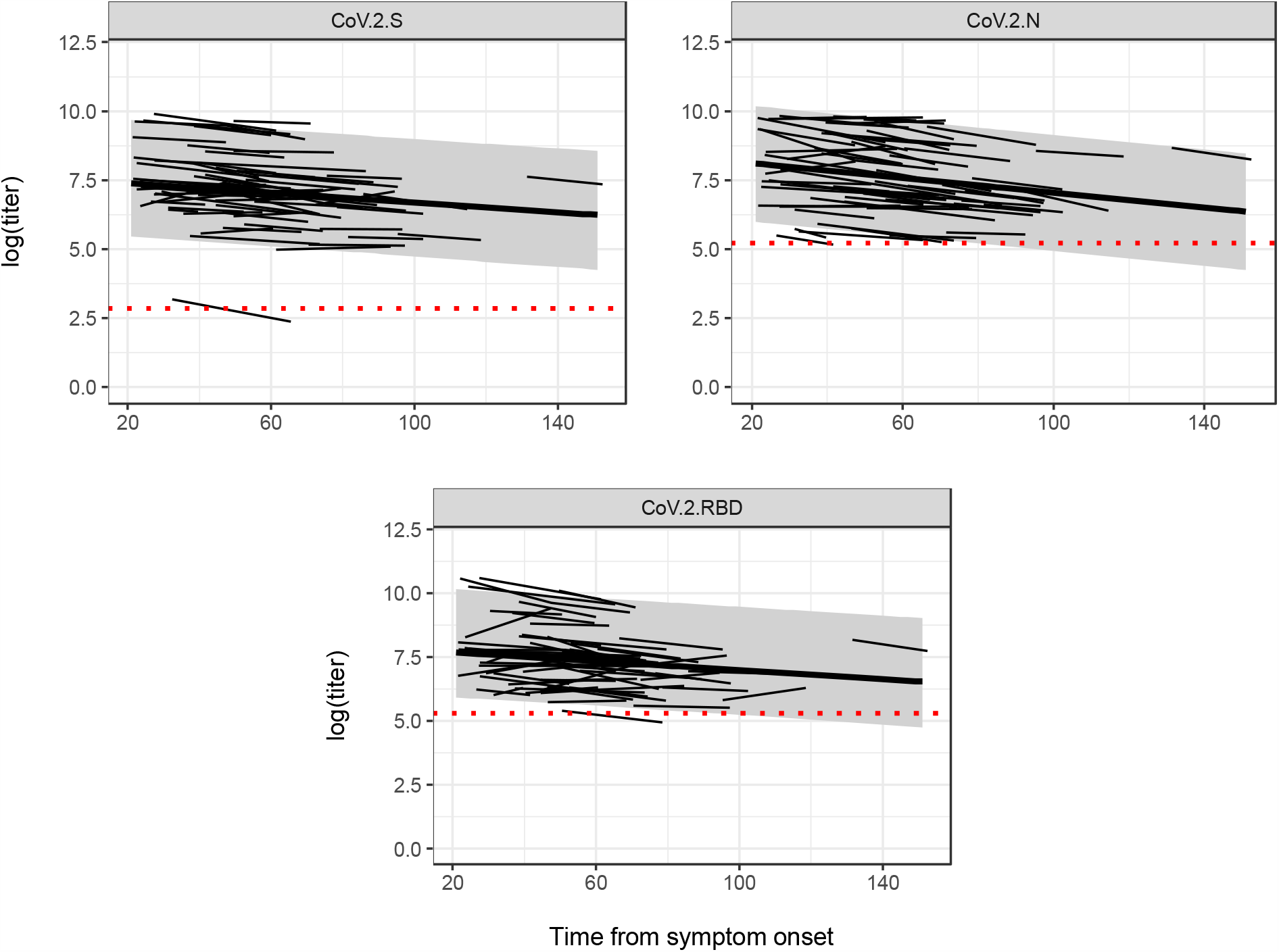
Bayesian log-linear model fits for the decay of the nucleoprotein (N) antibody, the trimeric spike (S) antibody and the receptor binding domain (RBD) antibody from 21 days following symptom onset. Confidence limits on the slope without individual random effects are plotted in dark grey, with individual effects light grey. Red dotted line represents the limit of seropositivity

**Figure 2.**
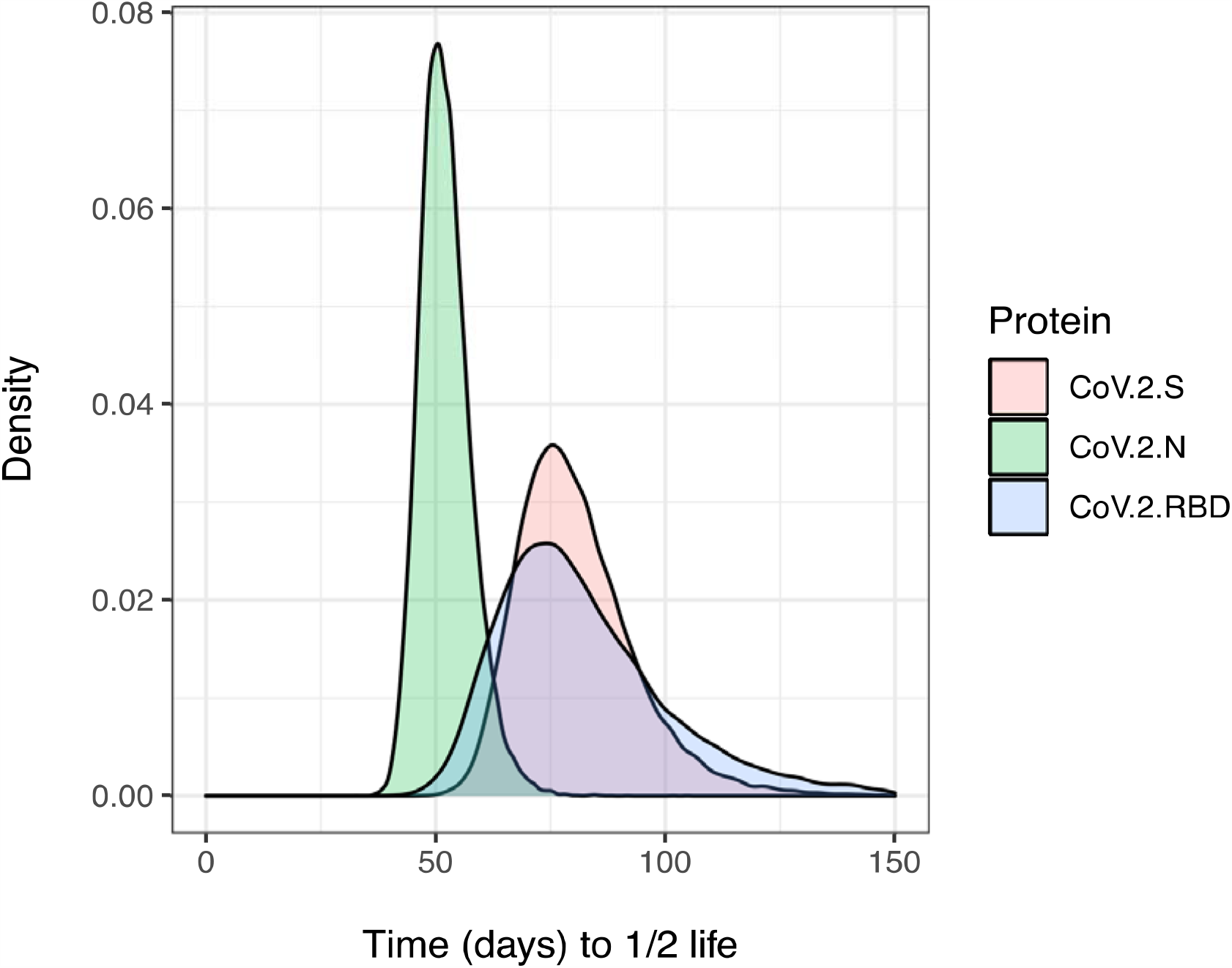
Posterior probability for the distribution of the half-life taken from the start of antibody decay at 21 days for the nucleoprotein (N) antibody (CoV.2.N green), the spike (S) protein antibody (Co.V.2.S red) and the RBD antibody (CoV.2.RBD blue).

**Figure 3.**
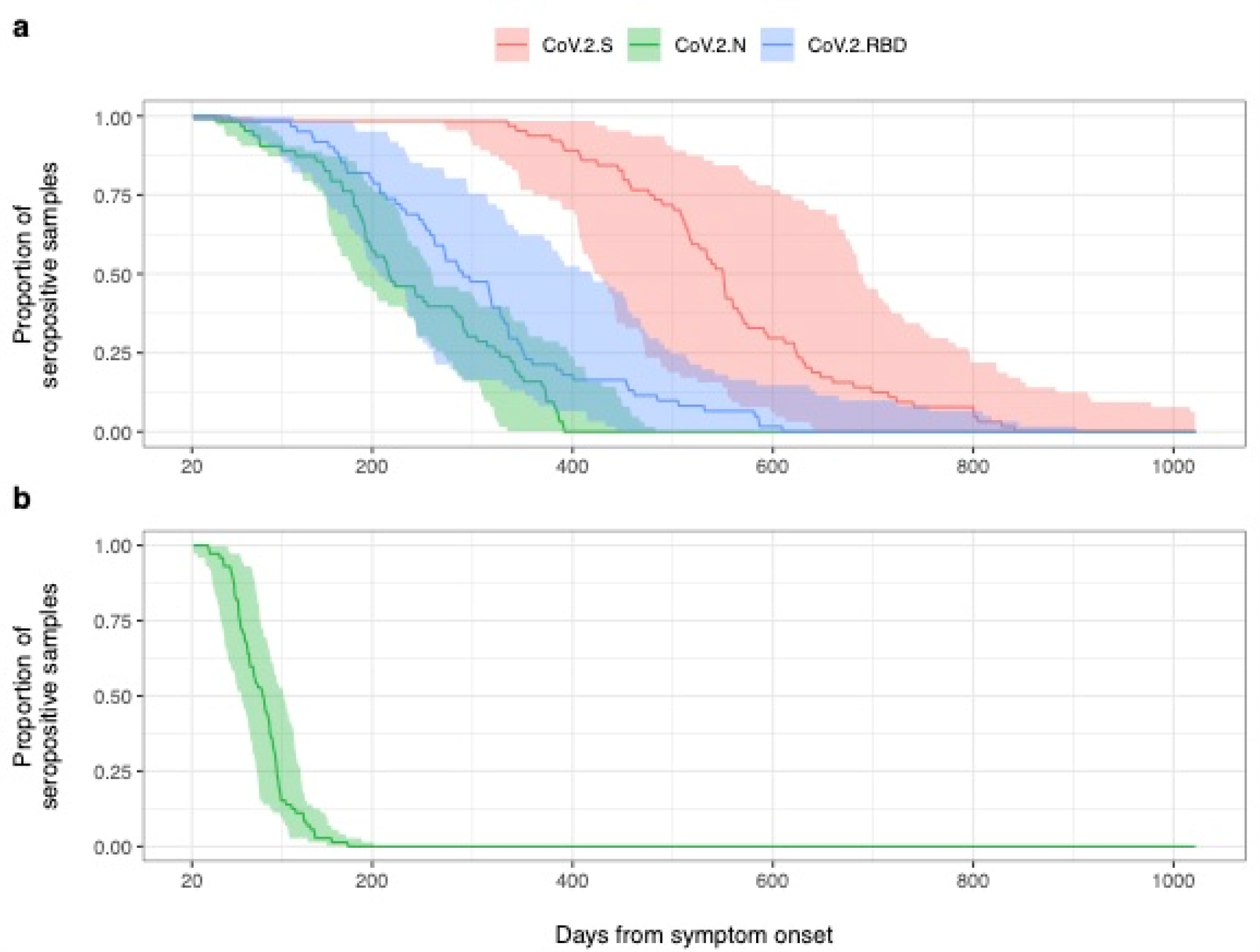
a) Time-to-negativity curve by the MSD assay for the nucleoprotein (N) antibody (green), the spike (S) antibody (red) and the receptor binding domain (RBD) antibody (blue). Time-to-negativity curve by the EDI assay for the nucleoprotein (N) antibody (green). *Note: The EDI assay only measures the N-antibody*.

### Half-life Estimations

The mean posterior probability of the slope of the N-antibody was twice the magnitude of the mean slopes of the S and RBD antibodies **(Supplementary Figure 4)**. This translated to a half-life of 52 days (95% CI 42-65) for the nucleoprotein antibody and a half-life of 81 (95% CI 61-111) and 83 days (95% CI 55-137) for the S- and RBD-antibody, respectively **(Figure 2)**.

**Figure 4.**
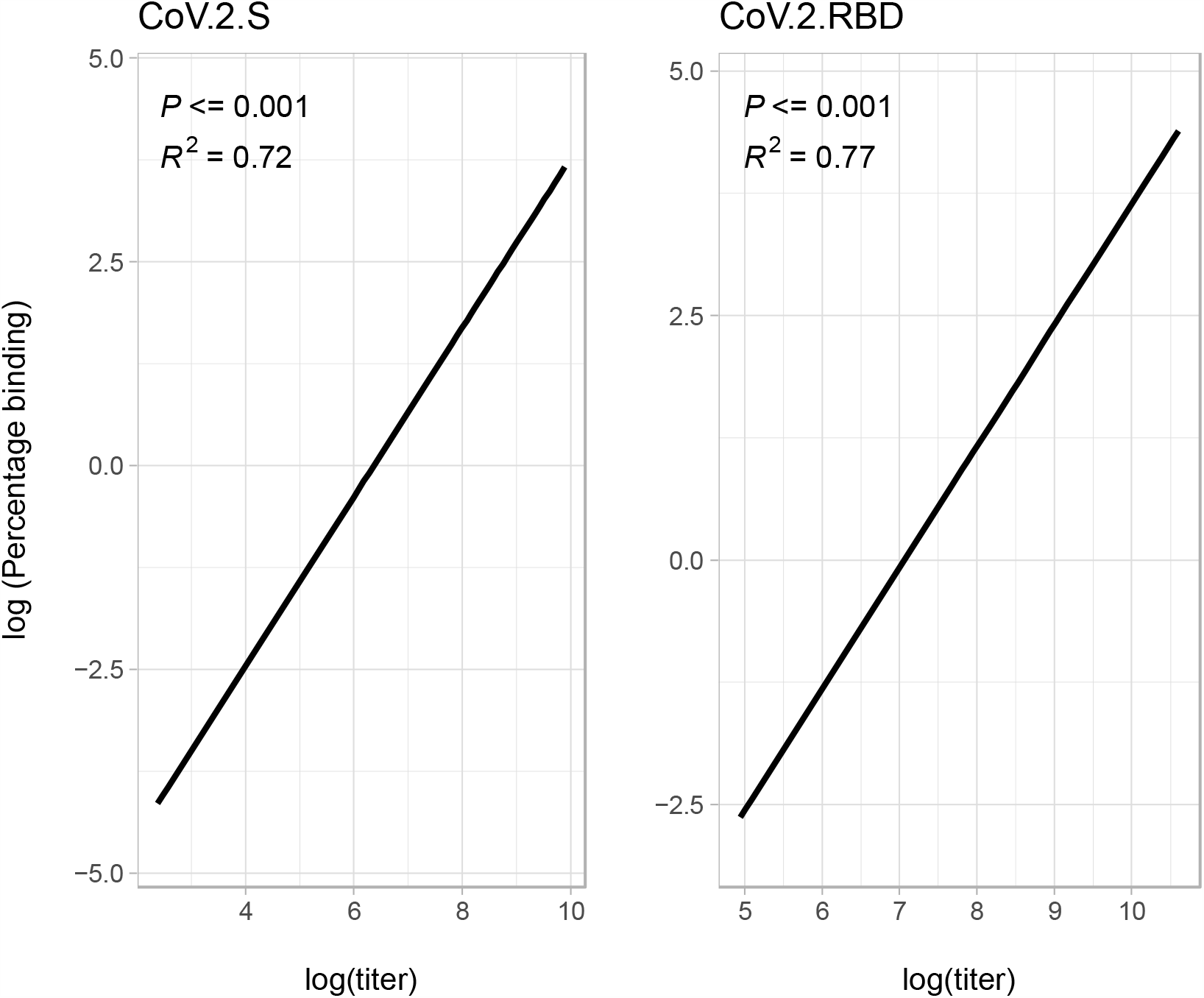
The correlation of SARS-CoV-2 Spike (S) and Receptor Binding Domain (RBD) antibody titers with ACE-2 receptor blocking.

### Time-to-Negativity

When these estimated rates were extrapolated to time-to-negativity **(Figure 3)** and time-to-undetectable curves **(Supplementary Figure 5)**, 50% of the seropositive population by the MSD assay were estimated to be negative to the N-antibody at 195 days (95% CI 163-236); by comparison, 100% of subjects were estimated to remain positive to the trimeric S-antibody at the same time point.

The time-to-negativity for 50% of the seropositive population for the RBD antibody was predicted to be 260 days (95% CI 180-418) and for the trimeric S antibody was 532 days (95% CI 418-667). However, the time-to-negativity was reduced to 67 days in the EDI assay (95% CI 47-91) days. **(Figure 3)**.

### Association of Demographic Variables with Antibody Titers

Both BAME participants and those >50 years were significantly associated with increased initial antibody titers. Neither co-morbidities, sex nor BMI were significantly associated with increased antibody titers. Interactions between age and time, sex and time and ethnicity and time were also examined and none were significant **(Supplementary Table 3 and Supplementary Figure 7)**.

### Competitive Binding Assay

There was a strong positive linear correlation between ACE-2 receptor blocking and both S-protein (R2=0.72, p<0.001) and RBD antibody titers (R2=0.77, p<0.001) **(Figure 4)**.

## Discussion

This prospective study of antibody responses following SARS-CoV-2 infection has estimated the mean half-life of the nucleoprotein (N) antibody to be only 52 days. Consequently, 50% of seropositive individuals will have a negative N-antibody test at 195 days after the start of antibody decay; by contrast 100% are predicted to have a positive test to the trimeric S-antibody at the same time point.

The half-life of the N-antibody was significantly shorter than that of the trimeric S- and RBD-antibodies with a mean of 81 and 84 days, respectively. These findings have immediate implications for SARS-CoV-2 diagnostic testing particularly widely used serological tests that target N-antibodies. Our findings are consistent with published estimates of SARS-CoV and MERS-CoV humoral dynamics following mild-to-moderate infection.(9, 20, 22, 26–28)

To date, no studies have comprehensively characterized the nature and duration of antibody responses to different SARS-CoV-2 epitopes, stratified by demographic factors and comorbidities. Long *et al* recently reported waning of SARS-CoV-2-specific IgG and neutralizing antibodies within 2-3 months following infection, decreasing by up to 76% and 81%, respectively. This was more pronounced following clinical disease, although 40% of asymptomatic patients (compared to 13% symptomatic) became seronegative in this early convalescent period.(10) Detailed antibody kinetics, however, were not reported. Asymptomatic or mildly symptomatic cohorts have also been largely overlooked, with most studies conducted on hospitalized patients. Our study addresses these gaps, facilitated by the MSD assay which enabled evaluation of absolute antibody titers to three major SARS-CoV-2 epitopes in parallel. Those participants over 50 years of age and BAME participants had higher initial antibody titers to N, S and RBD antigens. Although elderly patients are known to produce lower antibody responses to infection and vaccination,(29, 30) increased disease severity in this group may have influenced antibody response. The association between ethnicity and antibody titres needs to be further explored.

None of the seropositive healthcare workers identified in this study required hospitalization. This is important, given that only 1% of 20-30 year olds and 18% of >80 year olds diagnosed COVID-19 cases are hospitalized, making our study population representative of the majority of community SARS-CoV-2 infections.(31) Severe disease has been associated with higher antibody titers and a longer duration of antibody response following both SARS and MERS.(9, 20, 21, 32) When long-term follow-up studies of hospitalized SARS-CoV-2 patients are published, we therefore expect the half-life estimates to increase due to the increased severity of initial disease.

Viral neutralization assays remain the gold-standard *in vitro* correlate of protection and the lack of formal neutralization tests is a limitation of the study. ACE-2 receptor competition assays such as the MSD competitive binding assay have been shown to correlate well with formal viral neutralization assays(33). The ACE-2 competition assay used in this study had a strong positive linear correlation with both S-antibody titers (R^2^=0.72) and RBD-antibody titres (R^2^=0.77). Therefore, our observed decline in S-antibody titers is likely to be proportional to declines in functional receptor blocking activity.

Our negative exponential model of antibody decay assumed a constant rate of decay over time. Negative exponential models of antibody decay are frequently used to model antibody decay,(34, 35) and this assumption was supported by plotting the rate of decay over time **(Supplementary Materials Fig**.**2**).

Nevertheless, increased serial antibody measurements will enable us to test this hypothesis further and refine our half-life predictions accordingly. Whilst the severity of infection among our study participants is likely to be representative of community infection, our findings may be biased to healthcare workers. Moreover, a population of paediatric healthcare workers are more likely to be repeatedly exposed to seasonal CoV infection. We do not know to what extent this influences our measurements of the SARS-CoV-2 humoral response or our estimates of antibody decay. Recent studies have hypothesized that previous exposure to CoVs may confer some protection against SARS-CoV-2(36) and may need to be accounted for when modelling transmission or longevity dynamics.(37) Results are conflicting, however, and it remains unclear if any cross-reactivity is T-cell or antibody mediated.(38–41) Stratifying the SARS-CoV-2 humoral response by seasonal coronavirus titers will enable this potential bias to be understood.

Although all study participants had clinical symptoms that they attributed to COVID-19 **(Supplementary Table 1)** and at least one positive serological test, not all participants were diagnosed by PCR testing. Despite our formal performance evaluation of the MSD assay demonstrating a high sensitivity and specificity, it is theoretically possible that some of our paired positive samples arose from repeated false positive tests. Our estimates of the time-to-negative test for 50% of the population (t_neg50_) and the time-to-undetectable (t_u50_) are also dependent on the negative thresholds and lower limits of detection of the assay, respectively. Serological assays with very high positive-negative discrimination and low limits of detection will therefore detect antibody for longer. We repeated our analysis of N-antibody on the EDI™ assay which estimated an even lower t_neg50_ of 67 days (95% CI 47-91), suggesting that our estimates of the half-life of the N-antibody are robust between assays.

Protective immunity is a complex dynamic of host, environmental and viral factors. It can be difficult to disentangle the mechanisms underlying re-infection, including the role played by viral escape (i.e. genetic drift) versus weak or waning immune responses.(42) To date, no definitive quantitative or qualitative correlate-of-protection has been identified for SARS-CoV-2 infection, disease or onward transmission.(9, 43) This hampers our understanding of the functional and clinical significance of waning antibodies (particularly N-antibody) observed in this study. Recent findings from animal studies support the role of neutralizing antibodies as a correlate of immunity.(11, 16, 17) Similarly, a controlled human infection model of seasonal CoV 229E demonstrated waning of IgG-specific antibodies within 1 year, with subsequent reinfection of 6/9 individuals upon homologous viral challenge, albeit without clinical symptoms.(44) If reinfection does occur, severity of disease may therefore be attenuated, unless antibody-dependent disease enhancement by sub-neutralising antibody titers occurs.(42)

Furthermore, robust memory T-cell responses to specific SARS-CoV-2 peptides are elicited following infection; this also includes seronegative and/or pauci-symptomatic individuals.(15, 40, 41) As such, anamnestic cell-mediated immune responses may prevent clinically significant re-infection, in keeping with other viral pathogens such as measles and hepatitis A. Prospective evaluation of re-infection alongside T-cell, B-cell and mucosal IgA dynamics should therefore be an urgent priority. The long-term data generated by the Co-STARS study during the predicted future waves of the pandemic will be particularly informative.

In summary, this prospective cohort study of 137 longitudinally collected serological samples from 67 healthcare workers at Great Ormond Street Hospital has estimated that, in contrast to S-antibody, the half-life of IgG to N-antigen is only 52 days. Fifty percent will therefore have negative N-antibodies 195 days after the start of antibody decay. This has significant implications for sero-prevalence surveys that are based on the measurement of IgG to N-antigen in order to understand the spread of SARS-CoV-2 within the population.

Data of this kind directly informs public health policy and responses to future waves of the pandemic. Policymakers relying on these estimates at a population level may need to be cautious, allowing for possible underestimation of the true prevalence of previous SARS-CoV-2 infection and/or exposure. Conversely, the trimeric S-antibody correlated strongly with ACE-2 receptor blockade and persisted longer with 50% of seropositive participants estimated to still test positive at 532 days (t_1/2_ 81 days). This is reassuring for post-vaccine licensure surveillance studies, given that current vaccine candidates are based on recombinant or fragments of S-protein.(45) Our upper bound for any detectable antibodies (S, RBD or N) following mild-to-moderate infection was 1495 days.

## Materials and methods

### Study setting and design

Co-STARS is a 6-year single-centre, two-arm, prospective longitudinal cohort study of healthcare workers at a central London paediatric hospital (Great Ormond Street Hospital for Children). The study was approved to start by the United Kingdom NHS Health Research Authority on 29^th^ April 2020 and registered on ClinicalTrials.gov (NCT04380896). Informed consent was obtained from all participants. The Study Protocol and Supplementary Materials submitted with this paper include detailed methods, power calculations and the data analysis approach.

### Study participants

All hospital staff members ≥18 years of age were eligible for the study, provided they did not display symptoms consistent with SARS-CoV-2 infection at recruitment. If they had been symptomatic or had suspected contact with COVID-19 previously, at least 21 days were required to have passed prior to their baseline visit. Those significantly immunosuppressed(23) or those who had previously received blood products (including immunoglobulins or convalescent sera) since September 2019 were excluded from the study.

### Data Collection

After providing informed consent, participants undertook a detailed, standardised online questionnaire at study entry. This included socio-demographic factors, details of previous exposure to and symptomatic episodes consistent with COVID-19, any subsequent complications, previous SARS-CoV-2 diagnostic test results, past medical and contact history, and a comprehensive assessment of risk factors for exposure, susceptibility to infection and severe disease. Blood samples were also taken at baseline and each follow-up visit for determination of SARS-CoV-2 serology.

### Measurement of SARS-CoV-2 serum antibody and viral RNA by PCR

All serological samples were analysed by the Meso Scale Discovery (MSD) Chemiluminescent binding assay that detects and quantifies anti-SARS-CoV-2 IgG specific for trimeric S-protein, RBD and N-protein. All samples were run in parallel on the commercial ELISA EDI kit (Epitope Diagnostics Inc., California) that detects the anti-SARS-CoV-2 IgG to N-protein. The EDI kit did not quantify titers but did provide a positive, negative or equivocal test output with a ratio relative to the positive control.

An MSD® 96-well Custom Competition Assay designed to measure the inhibition of ACE-2 receptor binding to S or RBD by serum-derived antibody (MSD, Maryland) was run on 94 serial samples samples from 46 participants (two participants had 3 serial samples) in order to establish *in vitro* correlates of functional immunity. IgG levels for the MSD assay were expressed as arbitrary units calibrated against a set of reference sera distributed by the National Institute of Biological Standards and Control (Potters Bar, UK) under the auspices of the World Health Organisation. Assay qualification and performance were evaluated in our laboratories (see Supplementary Materials).

SARS-CoV-2 real-time reverse-transcriptase polymerase chain reaction (RT-PCR) targeting the N-gene was performed following RNA extraction, as previously described by colleagues at our laboratory.(24)

### Follow-Up Visits

All seropositive participants are being followed up monthly (ongoing) for repeat antibody testing for the first 6 months then 6-monthly thereafter. Seronegative participants will be followed up 6-monthly. At each follow-up visit, participants completed a shortened version of the baseline questionnaire, focussing on any recurrent COVID-19 exposure and/or symptoms.

### Study outcomes

The primary outcome of the study was to establish humoral dynamics following SARS-CoV-2 infection. Secondary outcomes measures included the incidence of SARS-CoV-2 re-infection, the dynamics of the cellular response, IgA dynamics and the clinical and demographic factors that are associated with the rate of antibody decay.

### Statistical analysis

Power calculations were based on log-linear regression using the pwr.f2.test function in R for general linear models. We assumed a study power of 80% and provided a study power for a variety of effect sizes and co-variates. We provided estimates for the study size required to detect effect sizes of ∼20% changes in log linear titers over 1 year with (with an alpha of 0.05) **(Supplementary Figure 1)**.

We used a Bayesian framework to model antibody decay from 21 days post infection as a negative exponential (log-linear) function of time using a random intercepts mixed effects model, with random effects at the individual level to adjust for the titer variation between patients. The 21-day starting point for antibody decay was chosen *a-priori* based on existing data of antibody responses to SARS-CoV-2.(25) Plotting the rate of decay against time supported our hypothesis that the rate of the negative exponential decay would remain constant over time (**Supplementary Figure 2)**.

Antibody titers were log-transformed and a linear model was fitted to the paired points using RSTAN in R (R: A language and environment for statistical computing. Foundation for Statistical Computing, Vienna, Austria. URL http://www.R-project.org/.).

Variability in antibody titers was modelled by introducing a random effects parameter at the level of the individual. Biologically important covariates that were hypothesized a-priori to influence antibody titers, including age, sex, BMI, comorbidities and ethnicity, were considered and the Deviance Information Criterion (DIC) was used to define the model of best fit to the data. The prior for the slope was drawn from a normal distribution (mean=0, variance=1), the intercept was a uniform distribution (min=0, max=10) while the random effects were taken from a normal distribution (mean=0, sigma_u) where sigma_u was estimated from the model.

The posterior probabilities of the slope were applied to infer the half-life distributions, using the following relationship between the half-life and the slope: t_1/2_ = ln(2)/*r* where *r* is the slope sampled from the posterior distribution and t_1/2_ is the half-life.

The time to test negativity for 50% of the population, t_neg50,_ was defined as the time for the initial antibody titers (measured from 21 days after symptom-onset) to decline to the negative threshold value for the assay. The confidence limits around the curves were derived by repeated sampling from the posterior distribution of the starting titers (including their random effect offset) combined with repeated sampling from the posterior slopes.

## Data Availability

All data will be freely available on request

## Acknowledgements

We would like to dedicate this article to the staff members who died of COVID-19 at Great Ormond Street Hospital during the first wave of the pandemic. We would also like to thank all the staff at Great Ormond Street Hospital who have taken part in the study. In addition, we are very grateful for all the hard work undertaken by the Great Ormond Street laboratory staff and the staff in the immunology laboratories both in the Camelia Botnar Laboratory and the Great Ormond Street Institute of Child Health who ensured that all the PCR tests and serological assays were completed in a timely manner. Finally, we would like to acknowledge the support of the Great Ormond Street Hospital Research & Development, Governance, Finance, Management, Estates, Operations and Communications departments.

